# Atrial Fibrillation Catheter Ablation among Cancer Patients: Utilization Trends and In-Hospital Outcomes

**DOI:** 10.1101/2023.11.13.23298490

**Authors:** Gilad Margolis, Ofir Goldhaber, Mark Kazatsker, Ofer Kobo, Ariel Roguin, Eran Leshem

## Abstract

**BACKGROUND:** Atrial fibrillation (AF) catheter ablation in cancer patients was evaluated in very few studies. We aimed to investigate trends of utilizations as well as in-hospital outcomes of AF catheter ablation procedures among cancer patients, in a large inpatient US registry.

**METHODS AND RESULTS:** Using the National Inpatient Sample (NIS) database, patients who underwent AF catheter ablations in the US between 2012 and 2019 were identified using ICD-9/10 codes. Sociodemographic, clinical data, in-hospital procedures and outcomes as well as in-hospital mortality and length-of-stay (LOS) were collected. Baseline characteristics and in-hospital outcomes were compared between patients with and without cancer. An estimated total of 67915 patients underwent AF catheter ablation between 2012-2019 in the US. Of them, 950 (1.4%) had cancer diagnosis. Compared with non-cancer patients, patients with cancer were older, had higher Charlson Comorbidity Index, as well as CHA2DS2-VASc and ATRIA bleeding indices scores.

Higher rate of total complications was observed in cancer patients (10.5% vs 7.9, p<0.001) driven mainly by more bleeding and infectious complications. LOS was also significantly longer in cancer patients (4.9 ± 5.8 vs. 2.7 ± 3.0 days, p<0.001). However, no significant differences in cardiac or neurological complications as well as in-hospital mortality rates were observed and were relatively low in both groups.

**CONCLUSIONS:** AF catheter ablation in cancer patients is associated with higher bleeding and infectious complication rates, but not with increased cardiac complications or in-hospital mortality rates in a nationwide, all-comer registry.

## INTRODUCTION

Atrial fibrillation (AF), is the most common sustained arrhythmia affecting 2-4% of the general population ^1^. Cancer patients have an increased risk for developing AF, varying according to cancer type and stage ^2,3^. Conversely, elevated cancer risk was observed among patients presenting with new-onset AF ^4^. The exact pathophysiological mechanisms leading to AF in cancer patients are not yet fully understood. Suggested underlying mechanisms include similar risk factor profile for both AF and cancer development, inflammatory and paraneoplastic processes, autonomic nervous system imbalance or less commonly direct metastatic invasion of the tumor to the heart and surrounding tissues ^5^. In addition, both medical and surgical cancer treatments were implicated as predisposing factors for AF development ^6,7^.

Rhythm control for symptomatic AF in cancer patients portends a therapeutic challenge. Significant drug-drug interactions with concomitant anticancer therapy may preclude antiarrhythmic drugs (AADs) use ^8^. Catheter ablation for AF is an effective and safe treatment strategy for symptomatic patients who do not benefit from AADs ^1^. However, to date, very few studies have evaluated AF catheter ablation among cancer patients ^9–14^. Current guidelines and expert opinion positions papers do not provide clear recommendations for AF rhythm control in cancer patients and specific recommendations for catheter ablation are lacking ^1,5,15,16^.

In this study we aimed to analyze trends of utilization and in-hospital outcomes of AF catheter ablation procedures among cancer patients using the US National In-Patient Sample (NIS) database.

## METHODS

### Data Source

The data were drawn from the NIS, the Healthcare Cost and Utilization Project, and the Agency for Healthcare Research and Quality. The NIS database includes only non-identified data; Therefore, this study was deemed exempt from institutional review by the local Human Research Committee. The NIS is the largest collection of all-payer data on inpatient hospitalizations in the United States. The data set represents an ≈20% stratified sample of all inpatient discharges from US hospitals ^17^. This information includes patient-and hospital-level factors such as patient demographic characteristics, primary and secondary diagnoses and procedures, comorbidities, length of stay (LOS), hospital region, hospital teaching status, hospital bed size, and cost of hospitalization. National estimates can be calculated using the patient-and hospital-level sampling weights that are provided by the Healthcare Cost and Utilization Project.

For the purpose of this study, we obtained data for the years 2012 to 2019. The International Classification of Diseases, Tenth Revision, Clinical Modification/Procedure Coding System (ICD-10-CM/ PCS) was fully implemented from the last quarter of 2015 and thereafter for reporting diagnoses and procedures in the NIS database during the study period. From 2012 to 2015 (3^rd^ quarter), hospitalizations were analyzed using ICD-9-CM/PCS coding system, and subsequently, with the ICD-10-CM/PCS coding system. For each index hospitalization, the database provides a principal discharge diagnosis and a maximum of 39 additional diagnoses, in addition to a maximum of 25 procedures.

### Study Population and Variables

We identified patients aged ≥18 years who had a primary diagnosis of atrial fibrillation using ICD-10-CM codes: I480, I481, I482, I4891 or ICD-9-CM code: 427.31 and also underwent Ablation using ICD-10-PCS codes: 02583ZZ, 02563ZZ, 02573ZZ, 025T3ZZ, 025S3ZZ or ICD-9-CM code:37.34. Using ICD-10-CM and ICD-9-CM codes (provided in detail in Table S1) we identified and excluded patients who had any of the following diagnoses: supraventricular tachycardia, atrioventricular nodal tachycardia, ventricular tachycardia, ventricular fibrillation, atrial flutter, ventricular or atrial premature beats, Wolf-Parkinson-White syndrome. To avoid inclusion of patients undergoing atrioventricular junction ablation only (as a “pace and ablate” strategy), we excluded patients with cardiac implantable electronic device (CIED) in-situ. We also excluded patients who underwent CIED implantation or open surgical ablation during their hospitalization. Similar data extraction methodology was previously utilized to identify patients undergoing AF ablation in the NIS registry^18,19^.

The following patient demographics were collected from the database: age, sex, and ethnicity. ICD-10-CM codes and ICD-9-CM codes (Table S1) were used to identify different comorbidities, including diabetes, hypertension, chronic heart failure, chronic kidney disease, obstructive sleep apnea, obesity, chronic pulmonary disease, anemia, hypertrophic cardiomyopathy, valvular heart disease.

For the purposes of calculating the Charlson–Deyo comorbidity index, additional comorbidities were identified from the database using ICD-10-CM codes and ICD-9-CM codes. The Charlson–Deyo comorbidity index is a modification of the Charlson comorbidity index, containing 19 comorbidity conditions with differential weights, with a total score ranging from 0 to 38 ^20–22^. Detailed information on Charlson–Deyo comorbidity index is provided in Table S2. Higher Charlson–Deyo comorbidity index scores indicate a greater burden of comorbid diseases and are associated with increased risk of death within 1 year after admission. The index has been used extensively in studies from administrative databases, with proven validity in predicting short-and long-term outcomes ^23–25^. In addition, CHA_2_DS_2_-VASc and ATRIA bleeding scores were calculated for each patient.

We obtained information on the presence of a known malignancy for each patient, based on ICD-10-CM codes and ICD-9-CM codes outlined in Table S3. Malignancies were categorized into two groups: hematologic malignancies and solid malignancies.

The primary outcome in this study was in-hospital complications including death. Secondary outcomes included sub-groups of complications, as well as LOS and total charges. Using ICD-10-CM/ PCS codes and ICD-9-CM/PCS codes (Table S1) the following in-hospital complications were identified: hemopericardium, tamponade, acute heart failure, cardiogenic shock, cardiac arrest, periprocedural hemorrhage/hematoma requiring transfusion, arteriovenous fistula, post procedure respiratory failure, periprocedural diaphragmatic disorder, periprocedural stroke and sepsis.

### Statistical Analysis

Frequencies and proportions of the different demographic, clinical, and hospital-related variables were calculated and weighted to reflect national estimates using discharge sample weights provided by the NIS ^24^. These estimates were compared according to cancer/non-cancer grouping using the pearson’s chi-square test and independent-samples t-test for categorical variables and continuous variables respectively. Generalized linear models were used to analyze annual trends. For all statistical analyses, we utilized SPSS® software version 23 (IBM Corp., Armonk, NY). A p-value < 0.05 was considered statistically significant.

## RESULTS

A total of 13,583 hospitalizations for AF ablation across the United States during the study period were included in the analysis. After implementation of the weighting method, these represented an estimated total of 67,915 hospitalizations for AF ablation. Most patients (61%) were men, and the mean age of the cohort was 64.4±10.9 years. Of the total estimated cohort, 950 patients (1.4%) had a diagnosis of cancer.

Baseline characteristics of the study population according to cancer/non-cancer grouping are presented in detail in Table 1. Older age, male gender, heart failure, chronic kidney disease and anemia were more prevalent in the cancer group. In addition, cancer patients had both higher CHA_2_DS_2_-VASc (2.72 ± 1.36 vs. 2.32 ± 1.45, p<0.001) and ATRIA bleeding scores (1.91 ± 1.86 vs. 1.21 ± 1.37, p<0.001; Table 1).

**Table 1:**
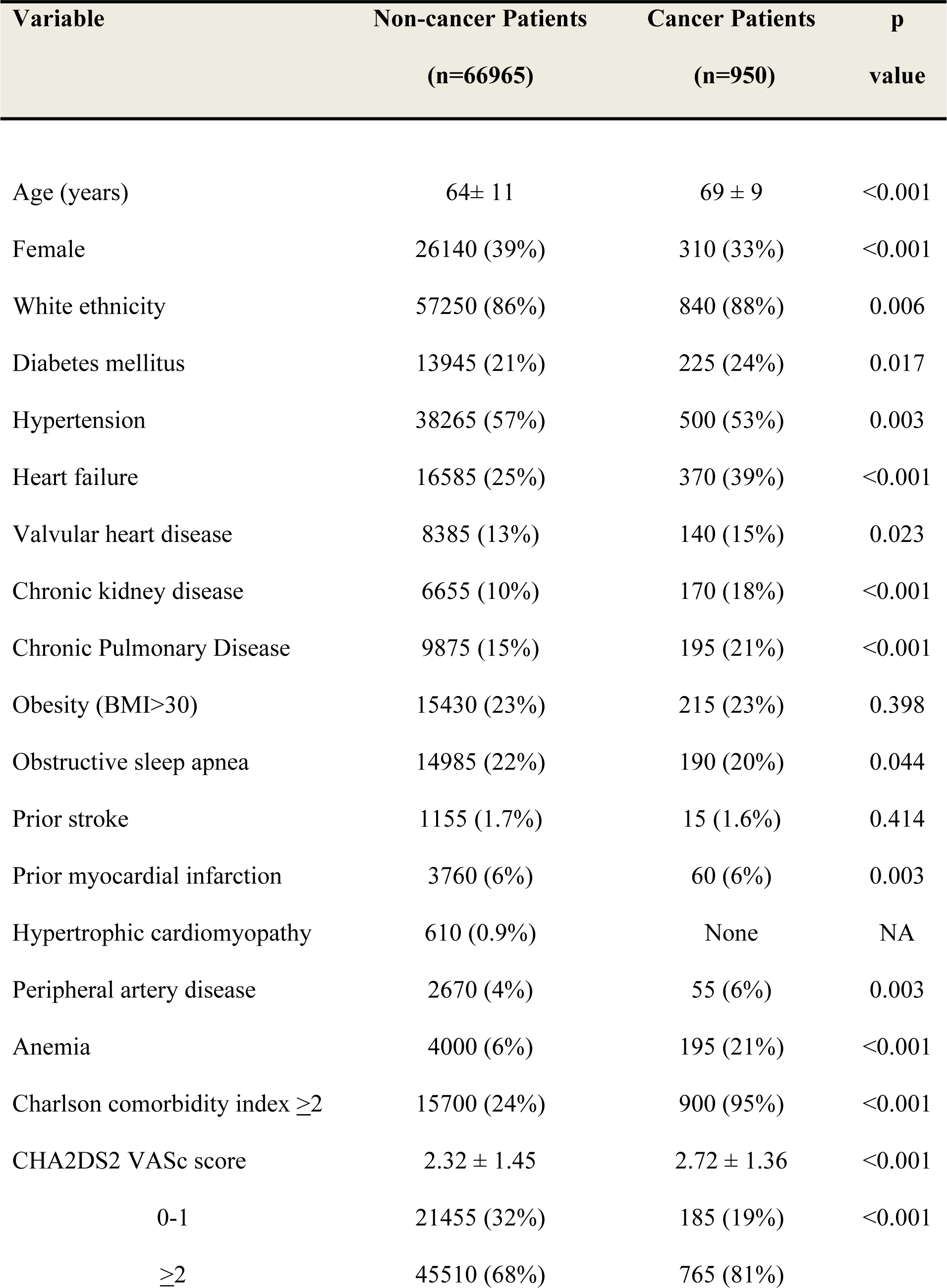
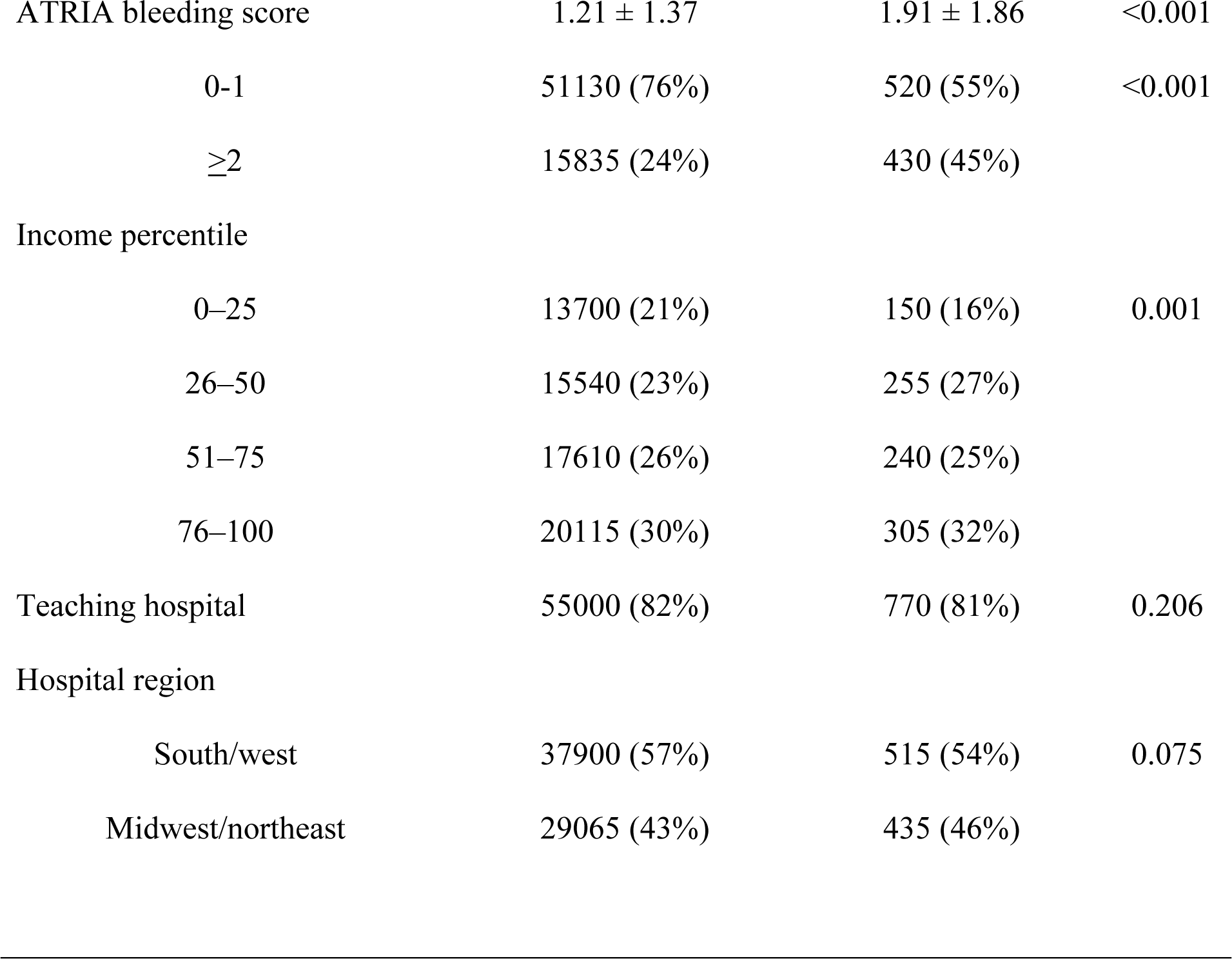
Baseline characteristics of patients undergoing AF catheter ablation.

| Variable | Non-cancer Patients<br>(n=66965) | Cancer Patients<br>(n=950) | p<br>value |
| --- | --- | --- | --- |
| Age (years) | 64± 11 | 69 ± 9 | <0.001 |
| Female | 26140 (39%) | 310 (33%) | <0.001 |
| White ethnicity | 57250 (86%) | 840 (88%) | 0.006 |
| Diabetes mellitus | 13945 (21%) | 225 (24%) | 0.017 |
| Hypertension | 38265 (57%) | 500 (53%) | 0.003 |
| Heart failure | 16585 (25%) | 370 (39%) | <0.001 |
| Valvular heart disease | 8385 (13%) | 140 (15%) | 0.023 |
| Chronic kidney disease | 6655 (10%) | 170 (18%) | <0.001 |
| Chronic Pulmonary Disease | 9875 (15%) | 195 (21%) | <0.001 |
| Obesity (BMI>30) | 15430 (23%) | 215 (23%) | 0.398 |
| Obstructive sleep apnea | 14985 (22%) | 190 (20%) | 0.044 |
| Prior stroke | 1155 (1.7%) | 15 (1.6%) | 0.414 |
| Prior myocardial infarction | 3760 (6%) | 60 (6%) | 0.003 |
| Hypertrophic cardiomyopathy | 610 (0.9%) | None | NA |
| Peripheral artery disease | 2670 (4%) | 55 (6%) | 0.003 |
| Anemia | 4000 (6%) | 195 (21%) | <0.001 |
| Charlson comorbidity index $\geq 2$ | 15700 (24%) | 900 (95%) | <0.001 |
| CHA2DS2 VASc score | 2.32 ± 1.45 | 2.72 ± 1.36 | <0.001 |
| 0-1 | 21455 (32%) | 185 (19%) | <0.001 |
| $\geq 2$ | 45510 (68%) | 765 (81%) | |
| ATRIA bleeding score | 1.21 ± 1.37 | 1.91 ± 1.86 | <0.001 |
| 0-1 | 51130 (76%) | 520 (55%) | <0.001 |
| ≥2 | 15835 (24%) | 430 (45%) |  |
| Income percentile |  |  |  |
| 0-25 | 13700 (21%) | 150 (16%) | 0.001 |
| 26-50 | 15540 (23%) | 255 (27%) |  |
| 51-75 | 17610 (26%) | 240 (25%) |  |
| 76-100 | 20115 (30%) | 305 (32%) |  |
| Teaching hospital | 55000 (82%) | 770 (81%) | 0.206 |
| Hospital region |  |  |  |
| South/west | 37900 (57%) | 515 (54%) | 0.075 |
| Midwest/northeast | 29065 (43%) | 435 (46%) |  |

Among cancer patients who underwent AF ablation during the study period, 495 (52%) had a solid malignancy, while 455 (48%) had a hematologic malignancy (Figure 1). Among solid malignancies, prostate cancer and lung cancer were most frequent, and leukemia was most frequent of the hematologic malignancies (Figure 1).

**Figure 1:**
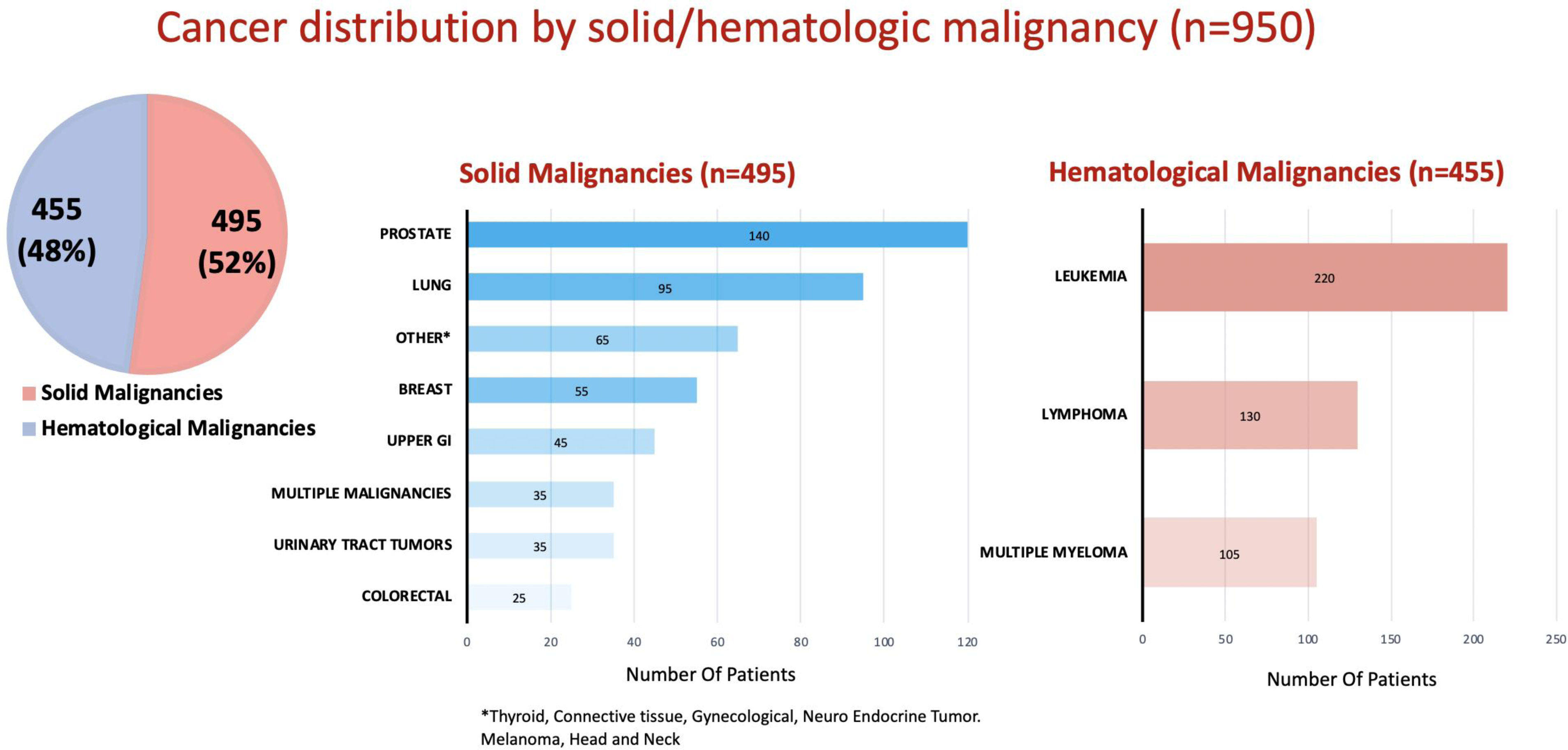
Cancer distribution by solid/hematologic malignancy NE= national estimate of hospitalizations.

The annual trend of AF catheter ablations for cancer patients gradually increased from 1.05% (n=115) in 2012 to 2.03% (n=170) in 2019 (p<0.001; Figure 2).

**Figure 2:**
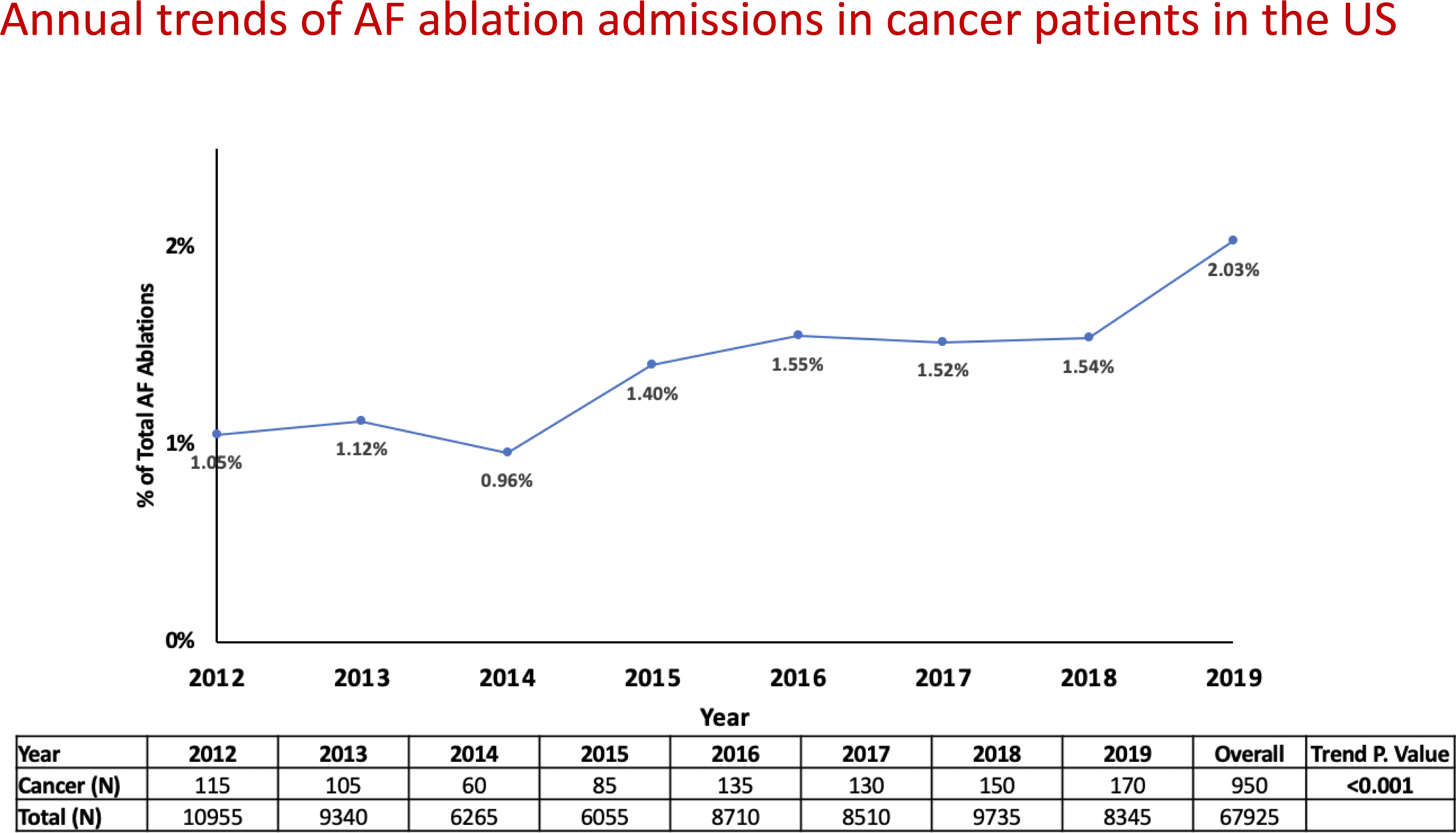
Annual trend of AF catheter ablations for cancer patients between 2012-2019.

### In-Hospital Course and Outcomes by Cancer/Non-cancer groups

In-hospital complication occurred in 5400 patients (8%) admitted for AF ablation. Compared with those without cancer, patients who had a cancer diagnosis had higher unadjusted rate of total in-hospital complications (10.5% vs 7.9%, p<0.001). The higher complications rate in the cancer group was driven by increased rate of bleeding (3.2% vs 1.8%. p=0.002) and infectious complications (2.1% vs 0.8%, p<0.001; Table 2). No significant differences were observed in cardiac complications, periprocedural stroke and in-hospital mortality rates, which were low in both groups (Table 2). The average LOS in the hospital was longer in cancer patients compared with non-cancer patients (4.91 ± 5.8 vs. 2.75 ± 3.05, Table 2).

**Table 2:** In-hospital diagnoses and procedures of patients undergoing AF catheter ablation with/ without cancer.

| Variable | Non-cancer Patients<br>(n=66965) | Cancer Patients<br>(n=950) | p<br>value |
| --- | --- | --- | --- |
| **Total complications** | 5300 (7.9%) | 100 (10.5%) | 0.002 |
| **Cardiac complications** |  |  |  |
| Cardiogenic shock | 80 (0.1%) | None | NA |
| Cardiac arrest | 100 (0.1%) | None | NA |
| Acute heart failure | 445 (0.7%) | ≤10 | NA |
| Hemopericardium/cardiac tamponade | 1085 (1.6%) | ≤10 | NA |
| **Vascular complications** |  |  |  |
| Hemorrhage/hematoma/blood transfusion | 1230 (1.8%) | 30 (3.2%) | 0.002 |
| Vascular injury | 1065 (1.6%) | 15 (1.6%) | 0.557 |
| **Respiratory complications** |  |  |  |
| Post-procedural respiratory failure/invasive ventilation >24 hrs | 1125 (1.6%) | 20 (2.1%) | 0.188 |
| Diaphragmatic disorders | 190 (0.2%) | None | NA |
| **Neurologic complications (stroke)** | 95 (0.1%) | ≤10 | NA |
| **Infectious complications (bacteremia, sepsis)** | 530 (0.8%) | 20 (2.1%) | <0.001 |
| **Length of stay** | $2.75 \pm 3.05$ | $4.91 \pm 5.8$ | $<0.001$ |
| **In-hospital mortality** | 165 (0.2%) | $\leq 10$ | NA |

Analysis of in-hospital complications by cancer type showed no significant difference in total complications rate between patients with solid or hematologic malignancies (10.1% vs 11%, p=0.36). However, patients with hematologic cancer had a significantly higher rate of bleeding complications, while those with solid cancer had higher rates of post-procedural respiratory failure and infectious complications (Figure 3).

**Figure 3:**
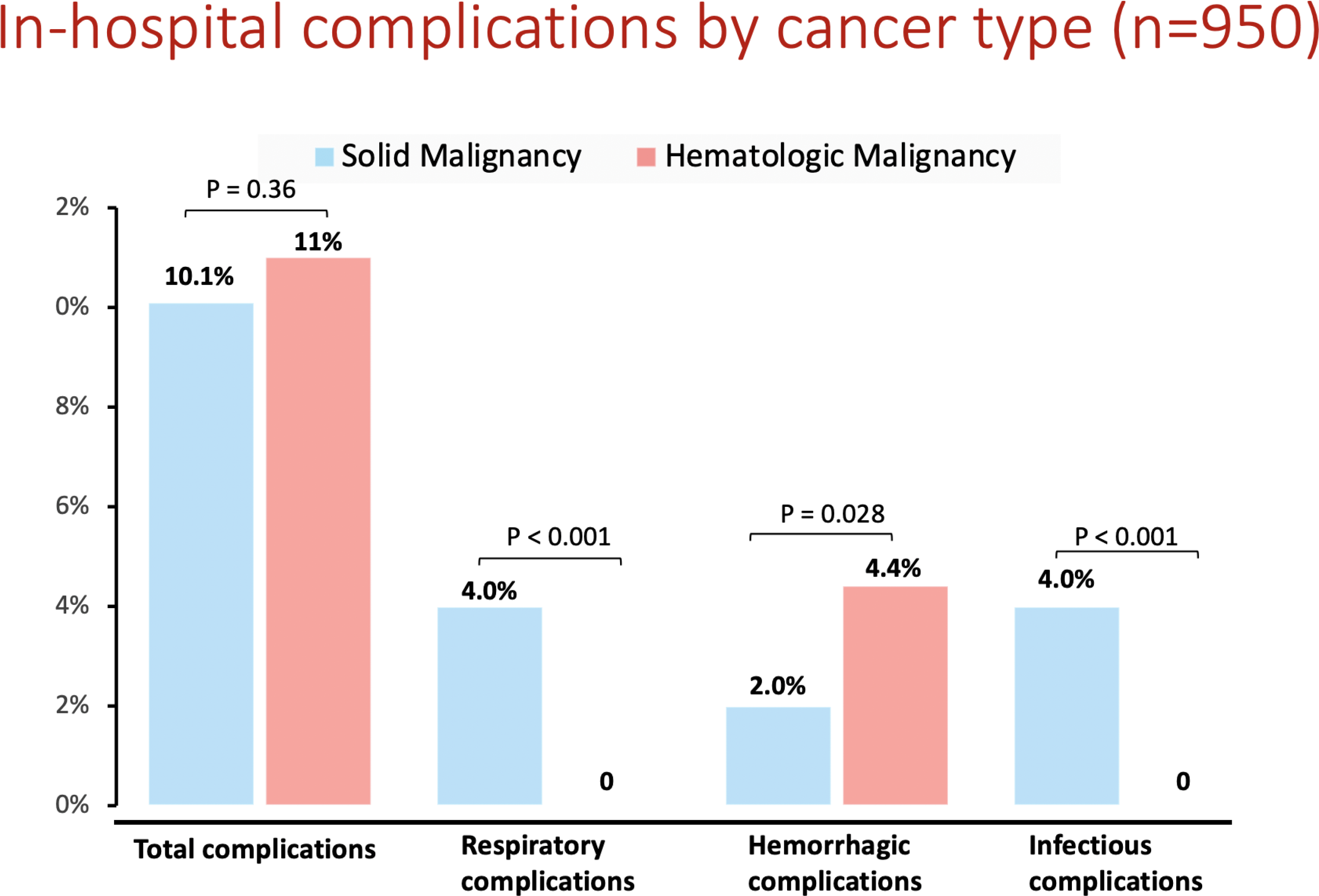
In-hospital complications by cancer type.

## DISCUSSION

Using data from the NIS, the largest all-payer inpatient database in the United States, we analyzed a weighted total of 67915 catheter ablation procedures for AF, between January 2012 and December 2019. Real-world nationwide data showed that 950 patients (1.4%) undergoing AF ablation had a cancer diagnosis. Annual trend analysis showed a gradual and significant increase of AF catheter ablations for cancer patients between 2012-2019. Cancer patients undergoing AF catheter ablation, were older, had more comorbidities and elevated thrombotic as well as bleeding risks, reflected by higher CHA_2_DS_2_-VASc and ATRIA bleeding indices, compared to their non-cancer counterparts. However, despite higher rate of total complications among cancer patients, driven by infectious and bleeding complications, no significant differences were observed in cardiac complications, periprocedural stroke or in-hospital mortality rates.

Current guidelines recommend pursuing rhythm control in symptomatic AF patients. ^1^ However, this may be challenging in cancer patients who also have AF. Maintaining sinus rhythm with anti-arrhythmic medications may involve significant drug-drug interactions with anti-cancer agents ^8^, There is scarce data on the efficacy of catheter ablation in this population. Nevertheless, observational data suggests that catheter ablation is equally effective in patients with or without a cancer history ^12,13^.

Very few studies assessed the safety of AF catheter ablation in cancer patients with discrepant results. Giustozzi et al. showed higher risk for clinically relevant periprocedural bleeding compared to non-cancer patients in 21 cancer patients who underwent AF catheter ablation.^11^ In two other studies, the frequency of periprocedural bleeding was similar between cancer and non-cancer patients ^12,13^. Of note, different periprocedural anticoagulation protocols were utilized in these studies, as no such protocol was ever verified in cancer patients. In our analysis, cancer patients had a higher rate of bleeding complications compared to non-cancer patients. We do not have data on the anticoagulation regimen given during and after the ablation in our cohort. Notably, the excess bleeding we observed was in patients with hematologic malignancies, which constituted almost half of patients, a higher proportion than in previous studies ^9–13^. In a retrospective analysis of patients undergoing percutaneous coronary interventions, leukemia diagnosis was associated with higher periprocedural bleeding complications ^26^. Whether hematologic cancer patients are at higher risk for bleeding complications in AF ablations needs to be evaluated in future studies.

Infectious outcomes were not reported in prior studies of AF ablation in cancer patients, who are potentially immunocompromised, due to both disease and treatment ^5^. In our study, we report for the first time a non-negligible rate of periprocedural infectious complications in cancer patients (2.1%), significantly higher than in non-cancer patients (0.5%). To date, there is no recommendation regarding periprocedural antibiotic prophylaxis for patients undergoing AF ablation ^27^. Guidelines for vascular and interventional radiology procedures recommend against antibiotic prophylaxis in patients undergoing cardiac procedures (e.g. coronary angioplasty), but give a IIb indication for antibiotic prophylaxis in patients undergoing solid tumor radiofrequency ablation, with the rationale being that thermal injury during this procedure may create a hospitable environment for bacteria ^28^. Whether cancer patients undergoing AF ablation need periprocedural antibiotic prophylaxis should be evaluated in future studies.

Recently, Thotamgari et al. ^14^ evaluated AF ablation procedure outcomes in 750 cancer patients identified in the NIS database between years 2016-2019. Utilizing propensity score matching technique, they reported a higher in-hospital mortality rate in cancer patients compared with non-cancer patients (2% vs. 0.7%). Importantly, patients with in-situ CIEDs were not excluded from their analysis. There are no specific ICD 9/10 codes for AV node ablation, so it is possible that such procedures were included in their analysis. As AV node ablation is usually reserved for the older and sicker patients, ^1,16^ the possibility of selection bias resulting in an unsound signal of excess mortality is conceivable. This could potentially explain their discrepant results with ours as well as with previous studies.^9–13^

### Study strengths and limitations

Several limitations should be acknowledged. The NIS database is a retrospective administrative database that contains discharge-level records and as such is susceptible to coding errors. Lack of information about patients’ cancer status in the NIS prevents us from distinguishing between patients with active cancer and patients with inactive disease, as well as concurrent anti-neoplastic treatment. We could only capture events that occurred in the same index hospitalization as the NIS does not include any follow-up data and all data is intentionally non-identified. The NIS database also does not include detailed information about patients’ clinical characteristics, medication, blood tests, and so on. These limitations are counterbalanced by the real-world, nationwide nature of the data, lack of selection bias, and absence of reporting bias introduced by selective publication of results from specialized centers. These results should not infer causation of periprocedural risk by malignancy, but merely present real world association that requires randomized trials for validation.

## CONCLUSIONS

The number of cancer patients undergoing AF catheter ablation procedures in the US increased steadily during the study period of 2012-2019. AF catheter ablation for cancer patients was associated with higher bleeding and infectious complication rates, but not with increased cardiac complications or in-hospital mortality in a nationwide, all-comer registry. These findings suggest that catheter ablation is a safe treatment modality for cancer patients who also experience AF.

## Abbreviations

AF: atrial fibrillation
AADs: antiarrhythmic drugs
CIED: cardiac implantable electronic device
Deyo-CCI: Deyo-Charlson Comorbidity Index
HCUP: Healthcare Cost and Utilization Project
ICD-10-CM/PCS: International Classification of Diseases, 10^th^ Revision, Clinical Modification/ *Procedure* Coding System
NIS: National Inpatient Sample
OR: odds ratio

## Data Availability

The national database data used for this study, analytic methods, and study materials will not be made available to other researchers for purposes of reproducing the results or replicating the procedure because of restrictions on the sharing of data in the Healthcare Cost and Utilization Project Data Use Agreement. The NIS database is publicly available for purchase, and the transparent and detailed methods that we have described make it possible for anyone who wishes to do so to replicate this study and reproduce our results

## Acknowledgments

None

## Source of Funding

None

## Disclosures

None.

### Supplemental Material

Tables S1-S3.

## Notes

### Competing Interest Statement

The authors have declared no competing interest.

### Author Declarations

The NIS database includes only non-identified data; Therefore, this study was deemed exempt from institutional review by the local Human Research Committee

